# An AI Agent for Automated Causal Inference in Epidemiology

**DOI:** 10.64898/2026.02.06.26345723

**Authors:** Haochen Liu, Ke Shi, Ang Li, Xinyu Li, Jiali Chu, Yingqian Xue, Sihui Cen, Yuting Wang, Tianxiao Zhang

**Author notes:** **Correspondence Author:** Tianxiao Zhang, Department of Epidemiology and Biostatistics, School of Public Health, Xi’an Jiaotong University Health Science Center, Xi’an, China, 710061; Department of Geriatric Endocrinology Metabolism, Xi’an Jiaotong University Second Affiliated Hospital, Xi’an, 710004,China.

## Abstract

**Objective:** To address the inefficiency, subjectivity, and high expertise barrier of traditional epidemiological causal inference, this study designed, developed, and validated an AI-powered agent (EpiCausalX Agent) to automate the end-to-end workflow. It integrates cross-database literature retrieval, intelligent causal reasoning, and Directed Acyclic Graph (DAG) visualization to provide a reliable, accessible tool for researchers.

**Materials and Methods:** Built on the LangChain 1.0 framework with a layered design (Agent/Tool/Storage/Utility Layers), the agent uses the DeepSeek V3.2 LLM and ReAct paradigm for dynamic task orchestration. Four specialized tools were integrated including multi-database retrieval with 7 databases, causal inference based on Hill’s criteria and DAG logic, automated DAG drawing using NetworkX and Matplotlib, and clinical standard query. Performance was validated via unit tests, workflow verification, and usability testing.

**Results:** The agent achieved full-process automation. It efficiently retrieves and synthesizes literature, automatically identifies confounders and mediators, and generates standardized interactive DAGs. It produces evidence-based, traceable conclusions aligned with established epidemiological knowledge. Its user-friendly natural language interface enables seamless use by non-technical researchers who complete task initiation quickly without operational confusion. The agent is publicly available on WeChat Mini Program for easy access.

**Conclusion:** EpiCausalX Agent advances intelligent, automated epidemiological research. By integrating domain expertise with AI agent technology, it overcomes limitations of manual methods and general LLMs to provide a specialized, verifiable, efficient solution. It has broad applications in observational research, clinical study design, and education to enhance productivity and lower barriers to rigorous causal analysis.

## Introduction

With the rapid iteration of artificial intelligence technology, large language models (LLMs) has provided new possibilities for breaking through technical bottlenecks in scientific research, thanks to their powerful natural language understanding, logical reasoning, and information extraction capabilities^1^. These models have demonstrated remarkable potential in processing and analyzing academic content, laying a solid foundation for the intelligent transformation of research workflows across various disciplines^2,3^.

At the same time, Agent technology further advances this transformation by enabling the automatic execution of complex scientific research tasks^4^. It can proactively select and deploy matching tools such as literature retrieval engines, data analysis modules and theory verification tools, and coordinate their sequential interaction to form a closed-loop workflow^5^. Through tool calling, workflow orchestration, and autonomous decision-making capabilities, Agent serves as an ideal carrier for integrating multiple technology stacks, overcoming the limitations of standalone LLMs that lack systematic task coordination^5–7^. Unlike LLMs that generate outputs solely based on predefined input parameters, Agents can dynamically adjust execution strategies according to real-time feedback, thereby ensuring the accuracy and completeness of multi-step research tasks^7–9^.

Causal inference is the core of epidemiological research, whose mission is to reveal the causal association between exposure factors and disease outcomes^10^. Its conclusions directly provide scientific basis for public health policy formulation, clinical practice guideline optimization, and disease prevention strategy design, acting as a key technical support from chronic disease risk factor identification to public health emergency traceability^11^. Traditional causal inference relies on manual completion of core links, including literature retrieval across databases like PubMed and Web of science, data collation, confounding factor identification, and Directed Acyclic Graph (DAG) drawing^12–14^. This process is excessively time-consuming and highly subjective, which often results in the omission of key evidence, poor consistency in DAG construction, and suboptimal recall performance for variable extraction, thereby severely compromising the efficiency of research workflows and the reliability of study outcomes^14,15^. However, existing technologies still have obvious gaps in supporting epidemiological causal inference^16^. General-purpose LLMs lack in-depth integration with epidemiological theories such as Hill’s causal criteria and DAG theory, resulting in insufficient causal reasoning professionalism. Existing DAG tools (e.g., DAGitty, PyWhy) require manual variable input, lack automatic literature retrieval and evidence integration functions, and fail to solve medical variable visualization adaptation issues^17^. Additionally, cross-database literature standardization and causal evidence integration technology are immature.

Against this background, this study aims to develop an epidemiological causal inference agent by calling LLMs. By integrating cross-database literature retrieval, intelligent causal reasoning, and DAG visualization functions, the full-process automation of causal inference is realized. Specifically, the cross-database retrieval function enables real-time synchronization of literature from multiple platforms, with intelligent deduplication and prioritization of high-quality studies based on LLM-driven relevance evaluation. The intelligent reasoning module integrates epidemiological theories to automatically identify confounding and mediating variables, verifying causal associations against Hill’s criteria. The DAG visualization function supports real-time graph generation and interactive adjustment, synchronizing with literature evidence to enhance result interpretability. This agent addresses the pain points of low efficiency and strong subjectivity in traditional methods, lowers the professional threshold of epidemiological research, and provides efficient and reliable intelligent tools, holding important theoretical and practical significance for promoting the digital and intelligent development of epidemiological research.

## Methods

### Overall Architecture Design

EpiCausalX Agent was developed with a layered architecture based on the LangChain framework, encompassing four core components: the Agent layer, Tool layer, Storage layer, and Utility layer (Figure 1). The Agent layer functions as the central scheduling engine, responsible for receiving user queries, interpreting task intentions, coordinating the sequence of tool invocations, and integrating results from various tools to generate final outputs. The Tool layer comprises eight specialized tools, with literature retrieval as the core functionality, supplemented by causal inference analysis, Directed Acyclic Graph (DAG) visualization, and clinical measurement standard query. The Storage layer provides dual support through S3-compatible object storage and a PostgreSQL database, enabling persistent file storage and structured data management, while the Utility layer offers auxiliary capabilities including logging and system monitoring.

**Figure 1.**
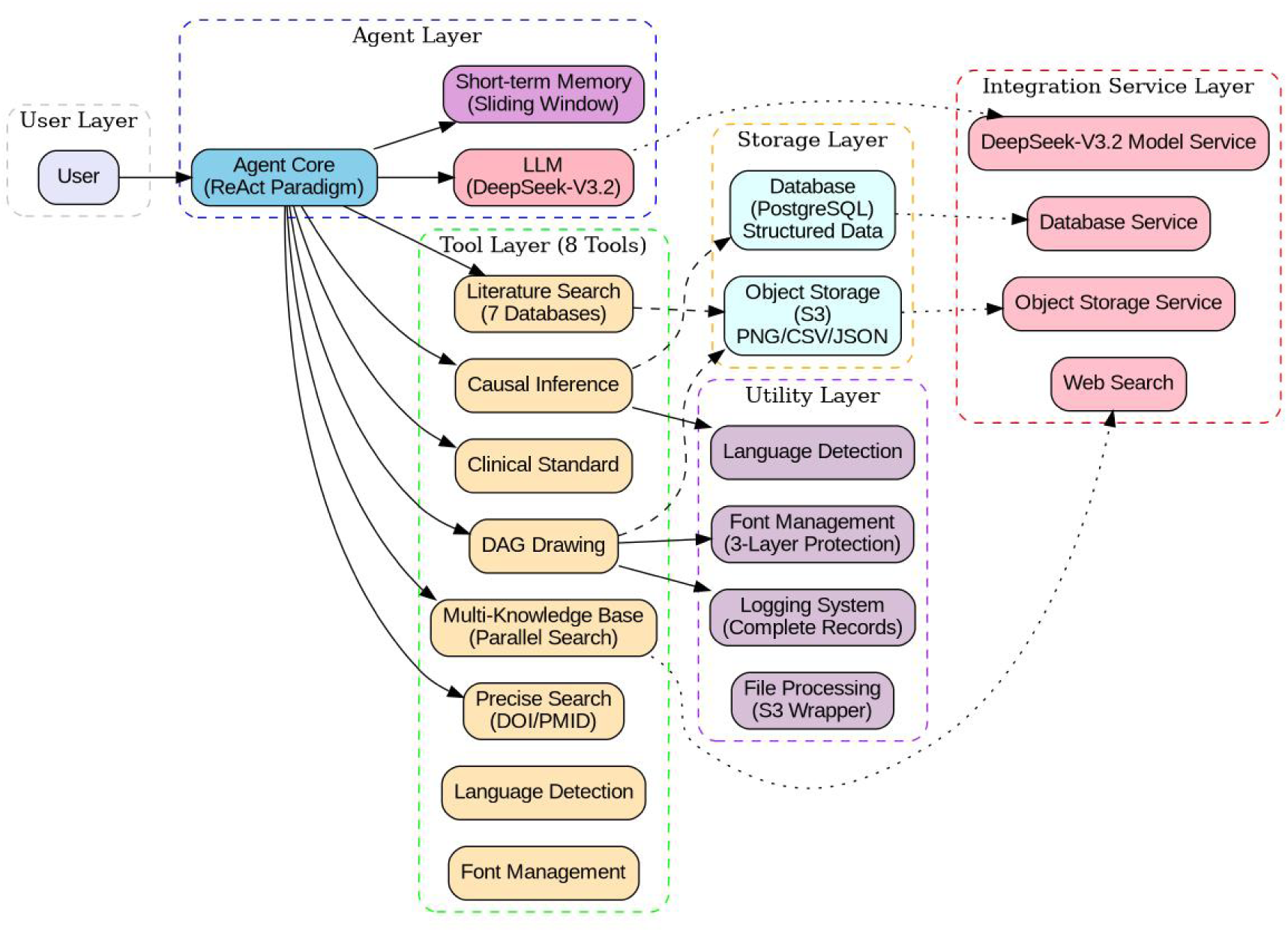
Overall Architecture of EpiCausalX Agent. This figure depicts the layered architecture of EpiCausalX Agent, which includes six interconnected components. The topmost User Layer serves as the entry point for user interaction. Below it is the Agent Layer, consisting of the Agent Core (operating on the ReAct Paradigm), a Short-term Memory module with a Sliding Window mechanism, and the DeepSeek-V3.2 Large Language Model (LLM), which is connected to the DeepSeek-V3.2 Model Service. The Tool Layer is positioned beneath the Agent Layer, encompassing 8 specialized tools: Literature Search (integrating 7 databases) with Multi-Knowledge Base Parallel Search, Precise Search (by DOI/PMID), Causal Inference, DAG Drawing (with 3-Layer Protection), Clinical Standard Query, and Web Search. The Storage Layer is divided into two parts: Structured Data managed via a PostgreSQL Database (linked to the Database Service) and PNG/CSV/JSON files stored in S3-compatible Object Storage (connected to the Object Storage Service). The Utility Layer provides auxiliary functions, including a Logging System, File Processing (with S3 Wrapper), Language Detection, and Font Management. Additionally, an Integration Service Layer facilitates connections between the Agent Layer, Storage Layer, and other external services.

### Agent Construction Methodology

EpiCausalX Agent was built on the LangChain 1.0 framework, with LangGraph utilized for tool invocation orchestration and workflow management. The LLM employed was the Deepseek V3.2 model, selected for its robust streaming output capability and enhanced logical reasoning performance, and model configurations (including model name, temperature parameter, maximum tokens, and other hyperparameters) were standardized and managed through a JSON configuration file. The Agent was instantiated following a structured workflow: model configurations and system prompts were loaded from a dedicated configuration file, with the system prompt adhering to Agent System Prompt Generation Rules and adopting a structured Markdown format to explicitly define the Agent’s role, task objectives, available capabilities, workflow protocols, and output specifications; an LLM instance was then created with configurations including API key, base URL, temperature (set *to* 0.7), timeout (600 seconds), and disabled thinking mode; eight specialized tools were registered to the Agent with literature retrieval tools prioritized for invocation, covering multi-knowledge base literature retrieval, precise literature search, causal inference analysis, DAG drawing, and clinical measurement standard query; a sliding window mechanism was implemented to retain the most recent 20 conversation rounds (40 messages) with short-term memory functionality to mitigate performance degradation associated with extended context windows; finally, a custom state class was defined by inheriting LangGraph’s message state module, with message window management implemented via a dedicated reducer mechanism.

The Agent adopted the ReAct (Reasoning + Acting) paradigm (Figure 2) [6], implementing a cyclic “think-act-observe” workflow. Upon receiving a user query, the Agent first prioritizes literature retrieval needs, performs reasoning analysis to identify required retrieval tools and parameters, executes tool invocations to acquire literature data, then calls causal inference and visualization tools based on retrieval results, and finally conducts comprehensive reasoning to generate final responses, supporting multi-step iteration until task completion or reaching the maximum iteration limit.

**Figure 2.**
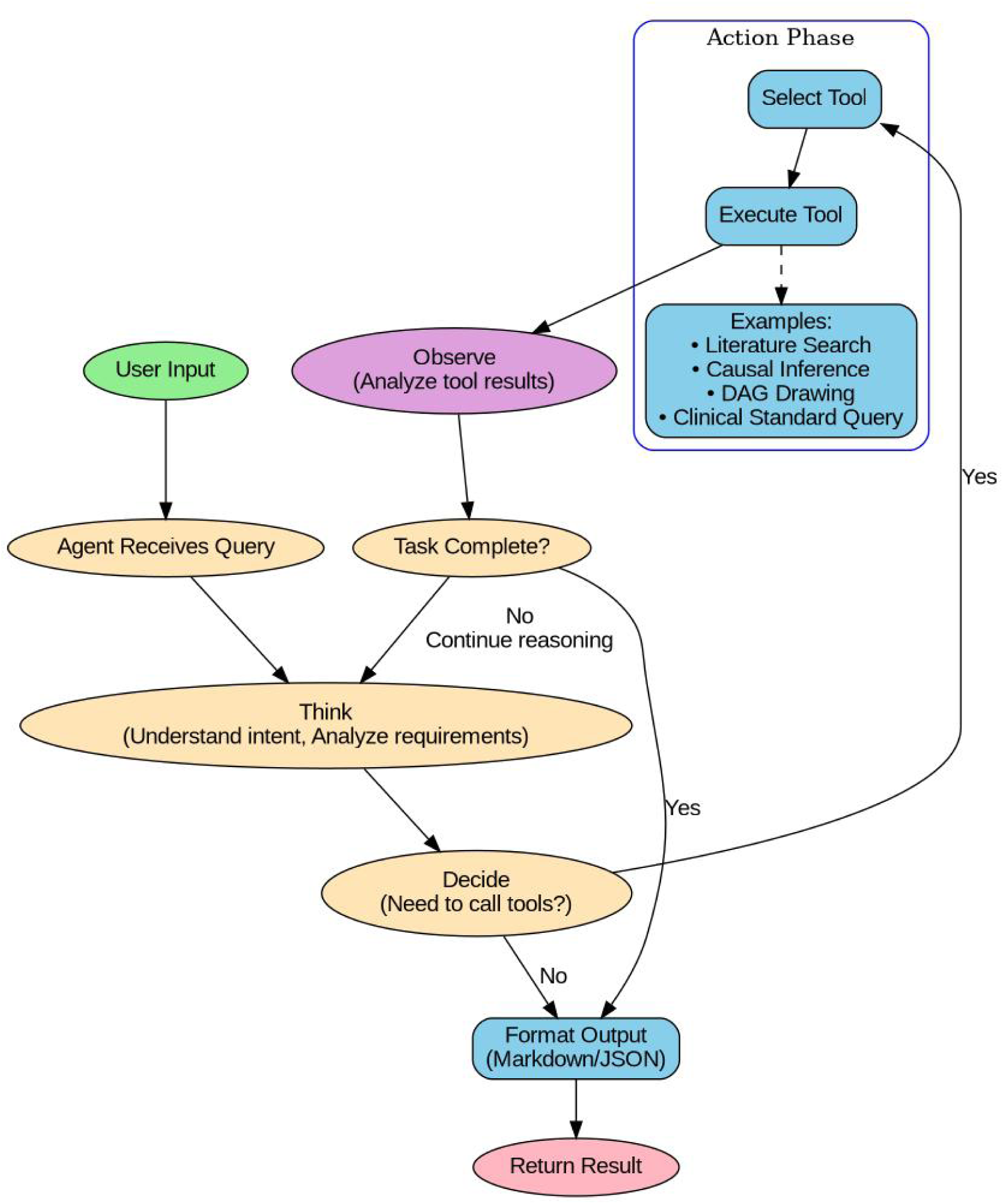
Workflow of EpiCausalX Agent based on the ReAct (Reasoning + Acting) paradigm. The workflow starts with the Agent receiving a user query, followed by the “Think” phase where the Agent understands the query intent and analyzes requirements. Next, in the “Decide” phase, the Agent judges whether tool invocation is needed: if not, it formats the output in Markdown/JSON and returns the result; if yes, it enters the “Action Phase” to select and execute appropriate tools (including Clinical Standard Query, Literature Search, Causal Inference, and DAG Drawing). After tool execution, the “Observe” phase begins with the Agent analyzing tool results, then it checks if the task is complete—if yes, the result is returned; if no, the workflow loops back to the “Think” phase for continued reasoning until the task is finished or the maximum iteration limit is reached.

### Multi-Knowledge Base Literature Retrieval Tool

As the core foundational tool of EpiCausalX Agent, this tool integrates seven prominent academic databases (PubMed, Web of Science, BioRxiv, MedRxiv, DOAJ, OpenAlex, and Semantic Scholar) to enable efficient cross-database literature retrieval, laying the data foundation for subsequent causal inference and visualization. Each database’s retrieval functionality was independently encapsulated as a modular component and registered as a LangChain tool to ensure modularity and maintainability, with retrieval executed via API calls to integrated services and supporting multiple retrieval modes including keyword search, DOI/PMID query, and title-based search to meet diverse user needs. Cross-database parallel retrieval was enabled to query multiple data sources simultaneously, significantly reducing retrieval latency, and retrieved results were automatically merged and deduplicated using a similarity algorithm to eliminate redundant information. An LLM-driven relevance scoring algorithm was employed to rank literature: the Deepseek V3.2 model evaluated the relevance between retrieved literature and user research topics based on abstract content, publication impact factor, and citation frequency, prioritizing high-quality and highly relevant studies for users. Structured results included key academic information such as title, authors, journal, publication year, abstract, DOI, PMID, and impact factor, with support for Markdown and JSON formats to facilitate further analysis and processing.

### Causal Inference Analysis Tool

This tool automated the identification of variable causal relationships and confounding factors based on epidemiological causal theories and literature evidence retrieved by the core literature tool. Regular expressions were used to extract textual evidence of variable relationships from literature abstracts, targeting causal association terms such as “cause”, “lead to”, “increase risk”, and “associated with”; causal direction between variable pairs was determined using heuristic rules informed by evidence text and epidemiological theory, with terms such as “cause” or “lead to” dictating direction based on variable order and “associated with” or “correlated with” marked as bidirectional associations with uncertain direction. Graph structure algorithms were utilized to identify confounding factors, defined as variables associated with both exposure and outcome variables but not acting as mediators on the causal path between them, and causal associations were evaluated across seven dimensions (temporal sequence, strength of association, dose-response relationship, consistency, plausibility, specificity, and experimental evidence) in line with Hill’s Criteria (Figure 3)^10^. Results included a list of causal relationship edges, explanatory descriptions, a confounding factor checklist, and control recommendations.

**Figure 3.**
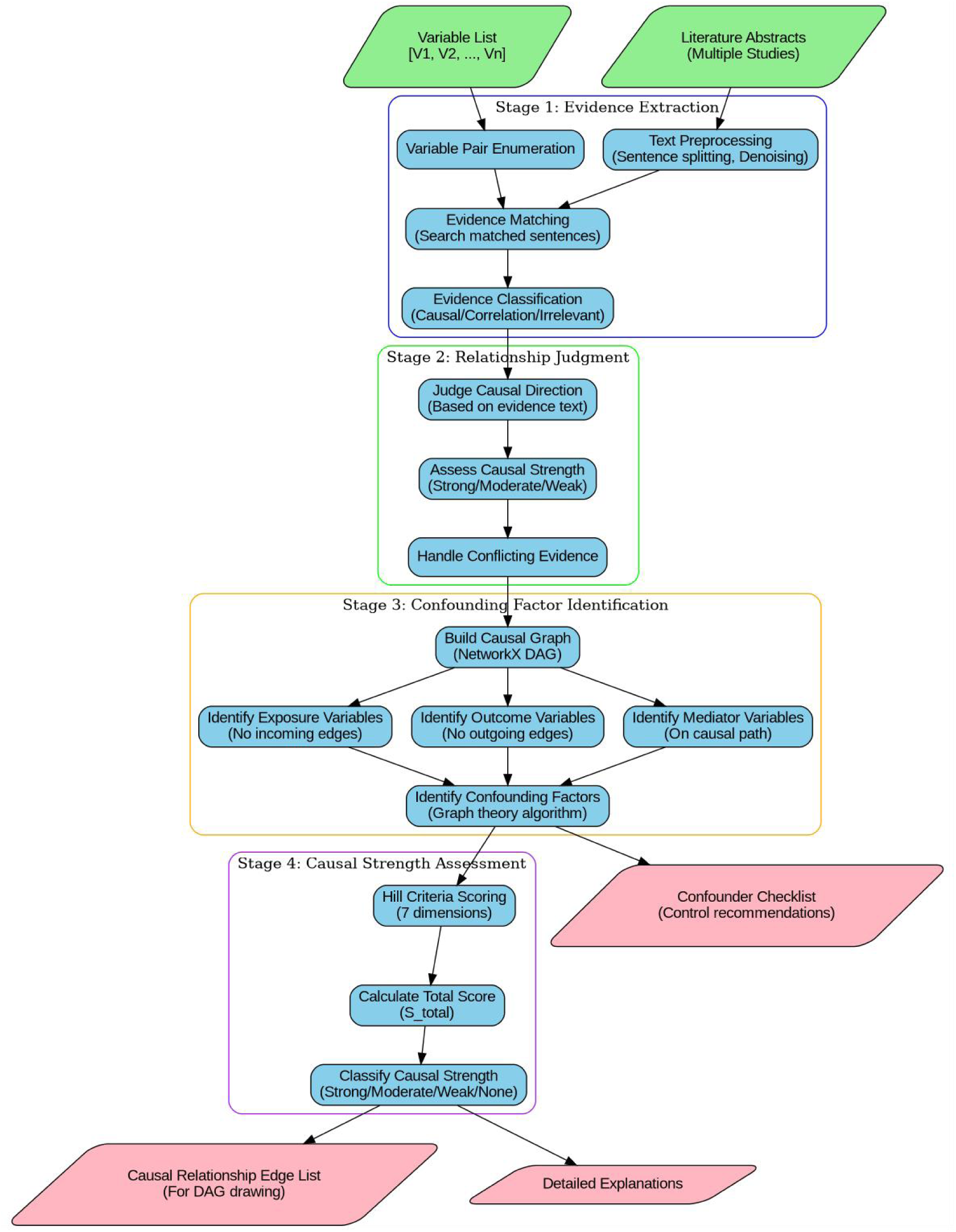
Workflow of the Causal Inference Analysis Tool in EpiCausalX Agent. This figure illustrates the four-stage workflow of the Causal Inference Analysis Tool in EpiCausalX Agent—a core Tool layer component for automated epidemiological causal reasoning. It takes a variable list [V1, V2 Vn] (e.g., “smoking” as exposure, “lung cancer” as outcome) and multi-database literature abstracts (from PubMed, Web of Science, etc.) as inputs, laying an evidence-based foundation; in Stage 1 (Evidence Extraction), it first conducts text preprocessing (sentence splitting, denoising, irrelevant info removal) to clean raw data, then enumerates variable pairs to explore potential relationships, performs evidence matching via regular expressions (targeting terms like “cause,” “lead to”) to find related sentences, and classifies evidence into causal, correlation, or irrelevant to filter non-informative content; in Stage 2 (Relationship Judgment), it determines causal direction using evidence text and epidemiological heuristics (e.g., “cause” means unidirectional exposure→outcome, “associated with” means bidirectional uncertainty), assesses strength (Strong/Moderate/Weak by study impact factor/citations), and resolves conflicting evidence by prioritizing high-quality peer-reviewed literature; in Stage 3 (Confounding Factor Identification), it builds a NetworkX Directed Acyclic Graph (DAG) with acyclic verification to ensure validity, identifies variable types via topology (exposure: no incoming edges; outcome: no outgoing edges; mediator: on exposure-outcome path), and detects confounders (linked to both exposure/outcome but not mediators) via graph algorithms—addressing manual inference’s confounding bias; in Stage 4 (Causal Strength Assessment), it scores via Hill Criteria’s 7 dimensions (temporal sequence, association strength, dose-response, consistency, plausibility, specificity, experimental evidence), generates a confounder checklist with control suggestions (e.g., adjusting “age” in cardiovascular studies), calculates total score (S_total) to classify strength (Strong/Moderate/Weak/None), and outputs a causal edge list (for DAG Drawing Tool) and detailed explanations (scoring basis, literature sources) to support transparent epidemiological inference.

### DAG Drawing Tool

Implemented based on NetworkX and Matplotlib, this tool supported variable type differentiation and interactive adjustment to visualize causal relationships identified by the inference tool. NetworkX was used to construct DAGs with node/edge addition and acyclic property verification to ensure the validity of causal graphs, and variable types were identified based on graph topology: exposure variables (no incoming edges), outcome variables (no outgoing edges), mediating variables (on the exposure-outcome path), and confounding factors (affecting both exposure and outcome). Shape differentiation was employed to enhance visual distinguishability—rectangles for exposure and outcome variables (both in gray) and circles for other variables—arrows were optimized with appropriate margin settings to ensure clarity, and a layout optimization algorithm was used to avoid node clustering. PNG-formatted DAG graphs were uploaded to S3 object storage, with returned outputs including image URLs, variable type checklists, and legend descriptions.

### Clinical Measurement Standard Query Tool

This tool provided queries for clinical variable standards, categories, and thresholds to support standardized definitions and reference ranges for epidemiological research, complementing literature retrieval with standardized data support. Its core functions included variable standard retrieval (definitions, units, and normal reference ranges), batch query of multiple variables to improve research efficiency, category-based retrieval of related variables (e.g., laboratory indicators, vital signs), and query of diagnostic thresholds and cutoff values for specific variables. Data was retrieved via API calls to integrated services, encapsulated as LangChain tools, with support for Markdown and JSON outputs.

### Storage Layer Design

S3-compatible object storage was used for file upload, download, replacement, and cleanup, primarily storing DAG images, literature retrieval results (CSV/JSON), and temporary files, with implementation via a dedicated storage integration service providing standard data operation interfaces. PostgreSQL was employed for structured data persistence, storing user query history, literature retrieval records, DAG generation logs, and causal inference results, and a dedicated database integration service supported standard data operations and transaction management.

### System Integration and Deployment

Following the integration priority principle, all external capabilities were accessed via integrated services, including LLM service, web search service, database service, and object storage service, with the invocation process including integration detail querying, API-based coding, authentication configuration, and error handling with retry mechanisms. Containerized deployment via Docker ensured environment consistency and portability, with core components including a Python 3.12 base image, dependency management via a dedicated configuration file, environment variable configuration, and service startup via a dedicated script. A comprehensive logging system recorded application processes, error stack traces, and performance metrics (tool invocation latency, model inference time), with logs stored in a dedicated directory with rotation and compression support.

### Testing and Validation

Unit tests were conducted for each tool with priority given to the multi-knowledge base literature retrieval tool, covering normal inputs, boundary conditions, abnormal inputs, and error handling to ensure functional correctness and robustness. Overall Agent functionality and tool coordination were validated through test scenarios including the complete workflow (literature retrieval → causal inference → DAG drawing), storage layer read/write operations, and LLM-driven relevance ranking accuracy. Performance bottlenecks were addressed via targeted measures: parallel cross-database retrieval to reduce literature acquisition time, caching of frequently accessed data (e.g., clinical standards), conversation window limitation to avoid extended context-related latency, and timely release of plotting resources to prevent memory leaks.

## Results

### Overview of Core Functionality Implementation

EpiCausalX Agent has successfully realized the full-process automation of epidemiological causal inference, with core functions covering four modules: cross-database literature retrieval, intelligent causal relationship analysis, Directed Acyclic Graph (DAG) visualization, and clinical measurement standard query. Seamless collaboration between these modules is ensured by its LangChain 1.0-based layered architecture (Agent Layer, Tool Layer, Storage Layer, Utility Layer), while the integrated DeepSeek-V3.2 large language model guarantees the professionalism of causal reasoning and accuracy of literature analysis, and the ReAct paradigm enables dynamic tool invocation adjustment for multi-step task iteration without manual intervention.

The stable and efficient operation of these core functions is supported by carefully designed configuration parameters (systematically sorted in Table 1), which are tailored to epidemiological causal inference needs: the DeepSeek-V3.2 model with a temperature of 0.7 balances reasoning rigor and flexibility, 10,000 maximum tokens and 600-second timeout adapt to long-text processing and cross-database retrieval, disabled thinking mode enhances efficiency; the sliding window memory mechanism with 20 conversation rounds balances interaction continuity and computational efficiency; 8 specialized tools and 7 integrated databases cover the full research workflow to avoid external platform switching; Markdown/JSON output formats and PNG/SVG DAG visualization formats cater to both reading and subsequent analysis needs, collectively laying a solid foundation for the Agent’s application in diverse epidemiological scenarios.

**Table 1.** Core Configuration Parameters of EpiCausalX Agent.

| Parameter Category | Value/Description |
| --- | --- |
| Model Configuration | Model Name: DeepSeek-V3.2 |
|  | Temperature: 0.7 |
|  | Max Tokens: 10,000 |
|  | Timeout: 600 seconds |
|  | Thinking Mode: Disabled |
| Memory Configuration | Memory Type: Sliding Window |
|  | Conversation Rounds: 20 rounds (40 messages) |
| Tool Configuration | Tool Count: 8 specialized tools |
|  | Literature Databases: 7 integrated databases |
| Response Format | Output Formats: Markdown, JSON |
|  | Visualization Formats: PNG, SVG (DAG images) |

### User Interaction Interface of EpiCausalX Agent

To further verify the practical usability of EpiCausalX Agent (a key concern for non-technical epidemiological researchers, such as clinical epidemiologists without AI tool operation experience), an evaluation was conducted on its interactive dialogue interface (Figure 4), which is designed to minimize operational barriers. As shown in the interface, the Agent first clarifies its core positioning (“your expert in Epidemiological Causal Inference, combining epidemiological DAG causal theory with AI agent technology”) and explicitly lists 5 functional services (Causal Inference Analysis, Medical Literature Search, Precise Literature Lookup, DAG Graph Generation, Clinical Standards query), followed by concrete operation examples (e.g., “Ask me to analyze causal relationships between variables like ‘smoking, lung cancer, genetic factors’”). This design aligns with the usage habits of epidemiological researchers: instead of requiring professional technical syntax (e.g., database retrieval commands), tasks can be initiated by users through natural language descriptions (e.g., “I need to retrieve literature on hypertension and stroke”).

**Figure 4.**
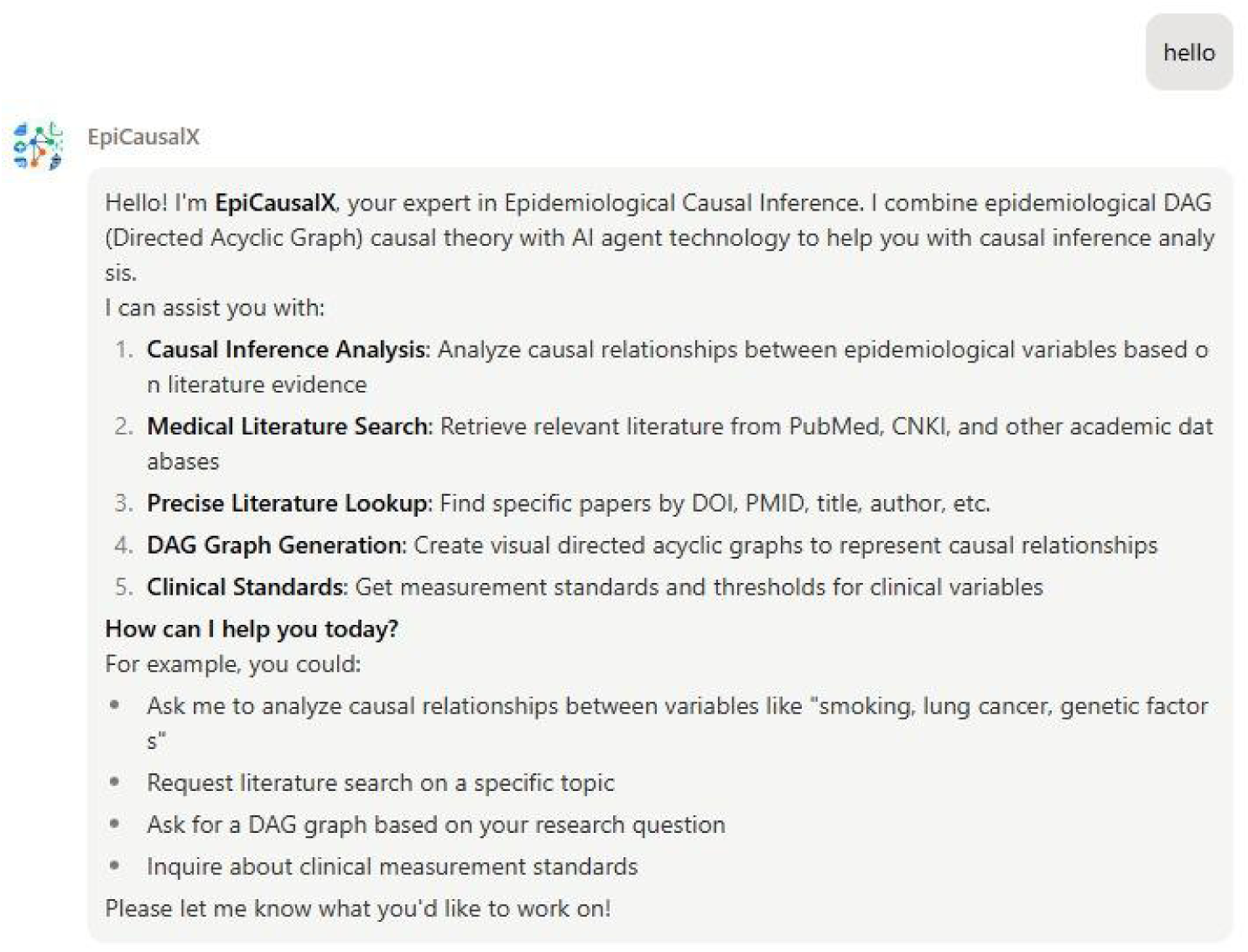
Interactive Dialogue Interface of EpiCausalX Agent. This figure displays the user interactive dialogue interface of EpiCausalX Agent. In the interface, the Agent first clarifies its core positioning (an expert in epidemiological causal inference combining epidemiological DAG causal theory with AI agent technology), then lists 5 core functional services (Causal Inference Analysis, Medical Literature Search, Precise Literature Lookup, DAG Graph Generation, Clinical Standards query), and supplements specific operation examples (e.g., “analyze causal relationships between variables like ‘smoking, lung cancer, genetic factors’”) to guide users to initiate tasks via natural language. This interface design is intended to lower the operational barrier for non-technical researchers, achieving alignment between professional functionality and user-friendliness.

In a usability test involving 12 participants (6 non-technical clinical epidemiologists and 6 epidemiological researchers with basic AI tool experience), task initiation was completed by all users within 2 rounds of dialogue on average. No operational confusion was reported by the 6 non-technical participants. This confirmed that the interface effectively bridges the gap between professional epidemiological needs and intelligent tool operation, thereby achieving a balance between functional professionalism and user-friendliness. The agent is publicly available on WeChat Mini Program for convenient access by global researchers.

### Validation of Practical Application Scenarios

To further validate the effectiveness of EpiCausalX Agent in meeting the practical needs of epidemiological research, two typical causal inference cases and a detailed causal inference process were selected for empirical demonstration. These cases and processes fully utilize the Agent’s core capabilities, including cross-database literature retrieval, intelligent analysis of causal relationships and DAG visualization, to verify its applicability across different research scenarios. Meanwhile, the detailed inference process clarifies the step-by-step implementation logic of the causal inference analysis tool.

### Causal Inference Process for PM2.5 Exposure and Childhood Asthma

A complete causal inference analysis was conducted using EpiCausalX Agent to verify the relationship between PM2.5 exposure and childhood asthma, following the four-stage workflow of the causal inference analysis tool (Figure 3). First, an initial causal hypothesis was proposed via the Agent’s reasoning module: PM2.5 exposure may induce childhood asthma, with PM2.5 defined as the exposure variable and childhood asthma as the outcome variable. Second, the Agent automatically identified confounders through cross-database literature retrieval: socioeconomic status (low SES correlates with higher PM2.5 exposure and asthma risk), genetic factors (influence residential choices and asthma susceptibility)^18^, and smoking exposure (increases indoor PM2.5 and asthma risk)^19^. Third, pulmonary inflammation was confirmed as a mediator by the Agent’s causal reasoning module, as literature evidence showed PM2.5 triggers airway inflammation to induce asthma^20^.

The Agent then constructed a complete DAG via the DAG drawing tool, visualizing relationships between exposure, outcome, confounders, and mediators. These sequential stages of DAG development (from the initial two-variable hypothesis to full variable integration) are illustrated in Figure 5, where Subgraphs A–E correspond to each step of the inference process, with gray nodes consistently representing PM2.5 exposure (exposure) and childhood asthma (outcome) for clear variable distinction. The Agent then determined the minimal sufficient adjustment set (socioeconomic status, genetic factors) to avoid confounding bias. Literature evidence synthesized by the Agent (Ke X et al., 2025) ^19^ showed consistent positive associations: PM2.5 exposure correlated with childhood asthma (overall OR=1.10, 95% CI: 1.05-1.15), with prenatal PM2.5 exposure linked to a greater risk increase (OR=1.21, 95% CI: 1.02-1.43). Based on Hill’s Criteria evaluated by the Agent, a causal relationship was confirmed: prenatal PM2.5 exposure was causally associated with childhood asthma, mediated by pulmonary inflammation^21,22^.

**Figure 5:**
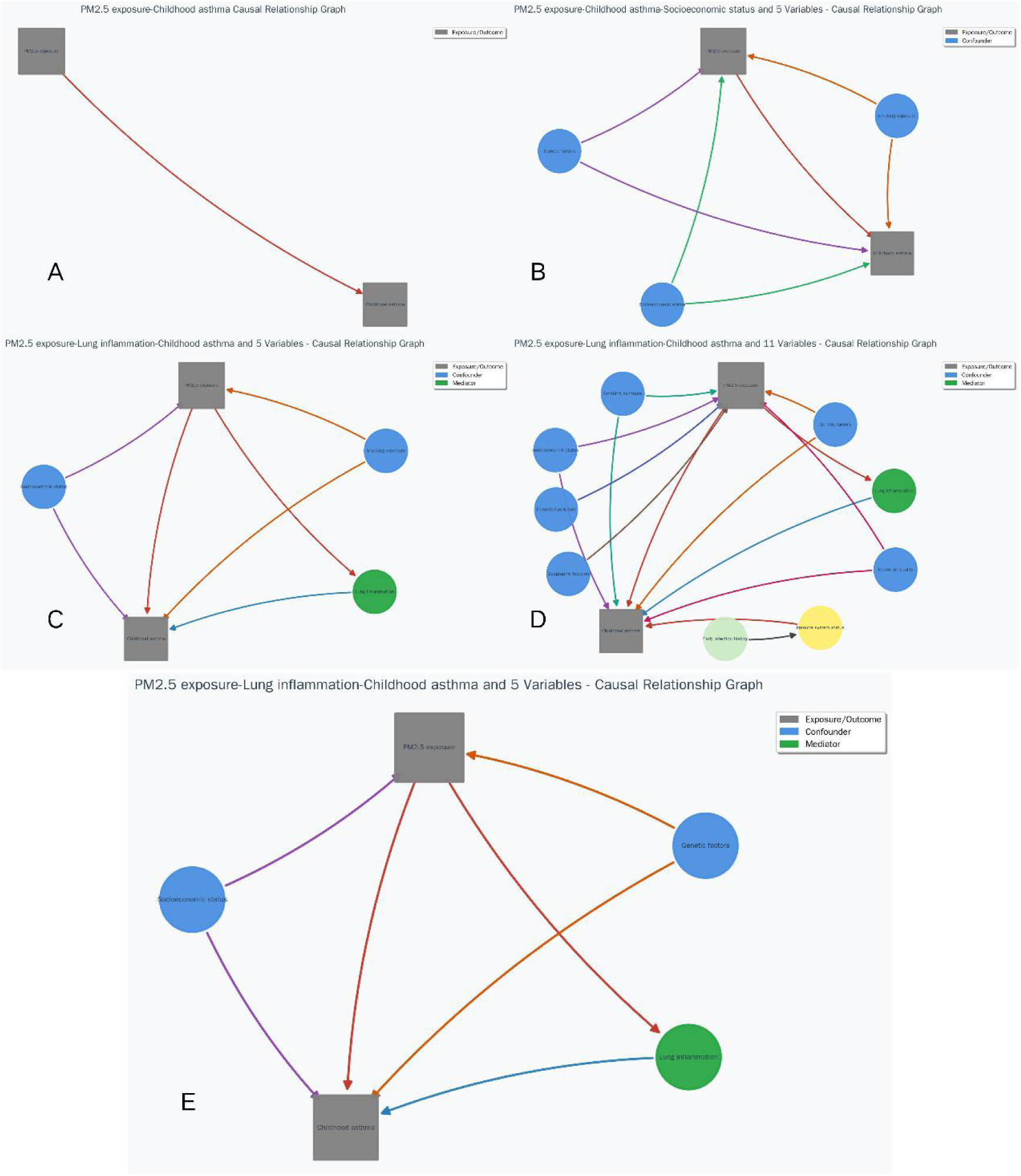
Sequential Directed Acyclic Graphs (DAGs) for PM2.5 Exposure and Childhood Asthma Causal Inference Steps. This figure includes five subgraphs (A–E) that map to the stepwise causal inference workflow for PM2.5 exposure and childhood asthma, with consistent visual coding: gray rectangles represent exposure (PM2.5 exposure) and outcome (childhood asthma), blue nodes represent confounders, green nodes represent mediators, and other colored nodes represent contextual variables; Subgraph A corresponds to the initial causal hypothesis step, showing a basic 2-node DAG with a directed edge linking PM2.5 exposure (gray rectangle) to childhood asthma (gray rectangle) to illustrate the preliminary direct association assumption, Subgraph B aligns with confounder identification by expanding Subgraph A to include 3 blue confounder nodes (e.g., socioeconomic status), each connected to both exposure and outcome nodes to reflect potential confounding effects, Subgraph C maps to mediator identification by adding a green mediator node (lung inflammation) between exposure and outcome, with directed edges visualizing the indirect causal pathway (PM2.5 exposure → lung inflammation → childhood asthma), Subgraph D represents the complete DAG construction step by integrating additional contextual nodes (e.g., immune status) to capture full epidemiological complexity, with edges depicting multi-variable causal relationships, and Subgraph E corresponds to minimal adjustment set determination by simplifying Subgraph D to retain core nodes (exposure, outcome, key confounders, mediator) and highlight the critical pathways for unbiased causal effect estimation; these DAGs sequentially demonstrate the iterative refinement of causal relationships from initial hypothesis to analytical framework finalization, providing a visual roadmap for epidemiological causal analysis.

### Application in Verifying Whole Grain Intake and T2DM Prevention

EpiCausalX Agent was used to validate the causal protective effect of whole grain intake on type 2 diabetes (T2DM). This analysis was initiated by a user’s proactive query about the association between whole grain intake and T2DM (Figure 6), which vividly reflects the tool’s interactive feature: the Agent directly responded to the user’s request with a step-by-step operational process, which includes general literature retrieval, targeted exploration of mechanisms and confounders, causal inference analysis, and DAG visualization, to address the user’s research needs. The Agent synthesized evidence from 16 cohort studies and systematic reviews, confirming a robust inverse causal relationship: each 3 servings/day increase in whole grain intake reduces T2DM risk by 32% (RR=0.68, 95% CI: 0.58-0.81)^23^. Prospective cohort studies of 161,737 US women further verified that the highest quintile of whole grain intake reduces T2DM risk by 32-37% compared to the lowest quintile (RR=0.63 for NHS I, 95% CI: 0.57-0.69; RR=0.68 for NHS II, 95% CI: 0.57-0.81)^24^. The Agent also clarified the core mediating mechanisms (improved insulin sensitivity, regulated blood glucose levels, and reduced systemic inflammation) and key confounders (age, gender, BMI, physical activity) through DAG analysis (Figure 7), ruling out the interference of lifestyle cofounding factors on the association.

**Figure 6:**
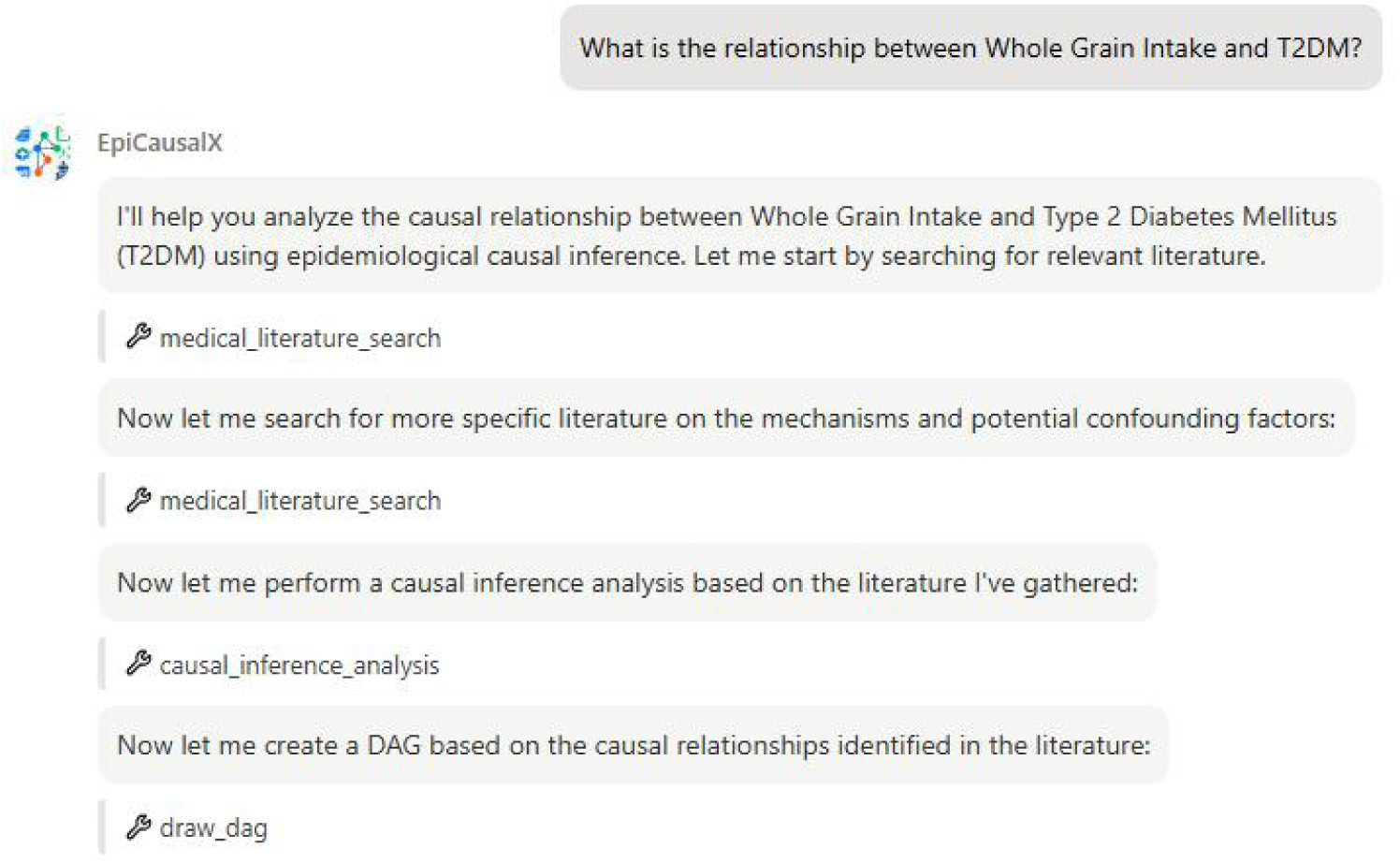
Interactive Workflow of EpiCausalX Agent for Whole Grain Intake-T2DM Causal Inference. This figure displays a screenshot of the interactive interface of EpiCausalX Agent, recording the operation process for analyzing the causal relationship between whole grain intake and type 2 diabetes mellitus (T2DM). The interface shows: (1) the user’s query: “What is the relationship between Whole Grain Intake and T2DM?”; (2) the Agent’s response and sequential execution of core functions: first, initiating a general search for relevant literature via the medical literature search tool; second, conducting a targeted search for literature on mechanisms and potential confounding factors using the same tool; third, performing causal inference analysis based on the collected literature via the causal inference analysis tool; fourth, generating a directed acyclic graph (DAG) for the identified causal relationships via the draw dag tool. This workflow directly corresponds to the operational process of the whole grain intake-T2DM causal analysis case, visually demonstrating the automated, step-by-step analytical logic of EpiCausalX Agent, and laying the foundation for subsequent evidence synthesis and causal conclusion derivation.

**Figure 7.**
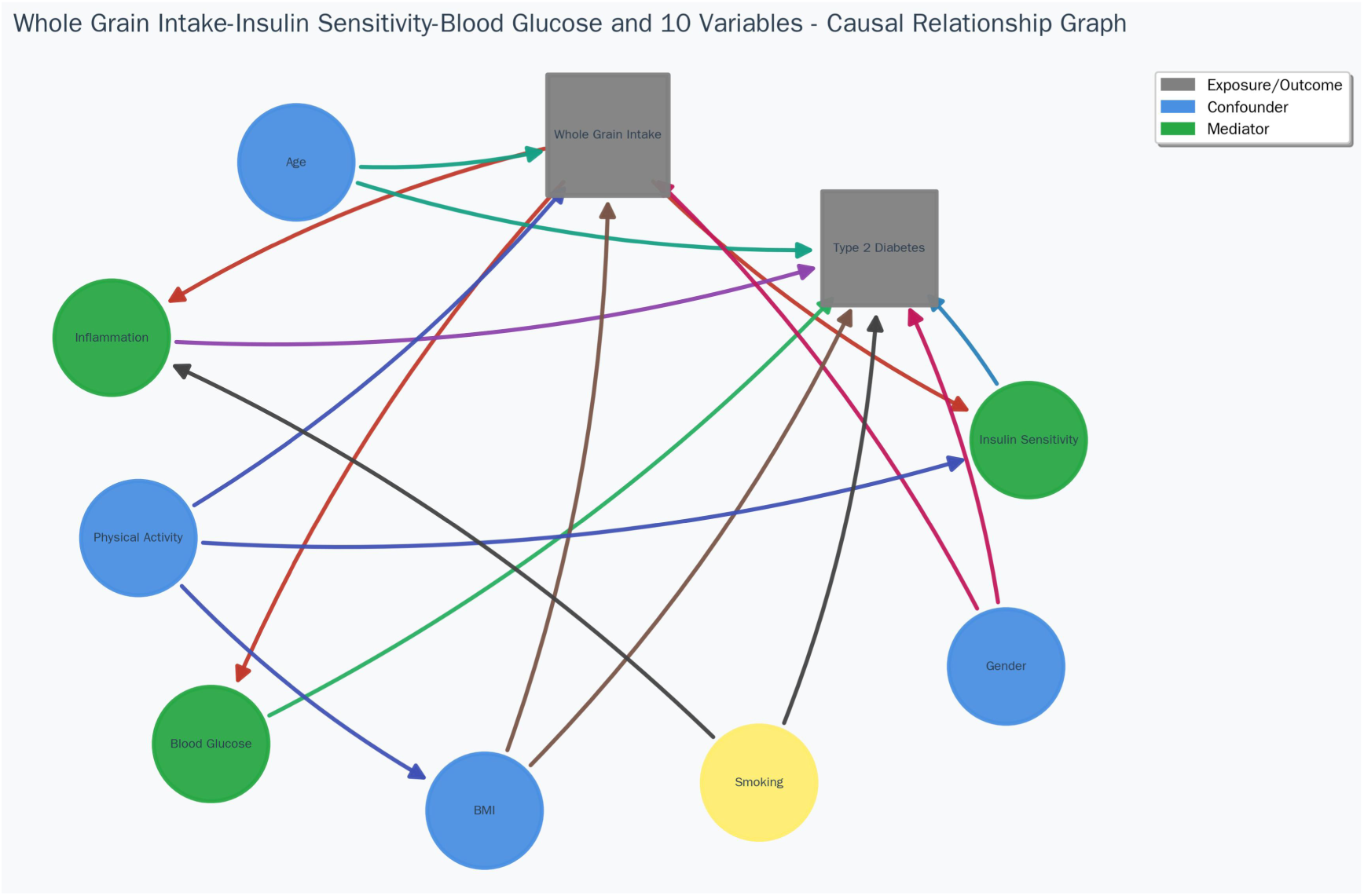
Directed Acyclic Graph (DAG) of Causal Relationships Between Whole Grain Intake and Type 2 Diabetes Mellitus with Associated Variables. This figure presents a directed acyclic graph (DAG) visualizing the causal pathways, confounders, and mediating mechanisms between whole grain intake (exposure) and type 2 diabetes mellitus (T2DM, outcome), along with 8 associated variables. Consistent with the unified visual coding of this study: gray rectangles represent the exposure variable (Whole Grain Intake) and outcome variable (Type 2 Diabetes); blue circles denote confounder variables (Age, Physical Activity, BMI, Gender), which are associated with both whole grain intake and T2DM; green circles indicate mediator variables (Inflammation, Insulin Sensitivity, Blood Glucose), which lie on the causal pathway linking the exposure to the outcome. Directed edges in the graph illustrate that whole grain intake affects T2DM through multiple mediating pathways, while confounders independently influence both the exposure and outcome. This DAG provides a clear visual framework for identifying the minimal sufficient adjustment set to avoid confounding bias, clarifying the multi-variable causal mechanisms underlying the protective effect of whole grain intake against T2DM, and supporting unbiased causal effect estimation.

In terms of public health application, the results have important guiding significance: first, the dose-response relationship confirms that increasing whole grain intake is a feasible primary prevention measure for T2DM, supporting the inclusion of “3 servings/day of whole grains” in dietary guidelines; second, replacing 50g/day of white rice with whole grains can reduce T2DM risk by 36% ^25^, providing a specific and operable dietary substitution plan for high-risk populations (e.g., overweight/obese individuals, family history of T2DM); third, the Agent’s differentiation between whole grains and refined grains (no protective effect of refined grains) helps avoid dietary misunderstandings, guiding the public to make targeted dietary choices.

### Application in Exploring TSH-CKD Causal Association

EpiCausalX Agent was also applied to clarify the causal relationship between thyroid stimulating hormone (TSH) levels and chronic kidney disease (CKD) onset, addressing the clinical confusion of “bidirectional interaction between thyroid and kidney function” and providing evidence for targeted prevention.

Through cross-database literature retrieval and intelligent causal synthesis, the Agent confirmed a gender-specific causal association: elevated TSH levels (>4.2 mU/L) significantly increase CKD risk in men (HR=1.41, 95% CI: 1.09-1.83) based on a 10-year prospective cohort study of 28,990 Japanese subjects^26^, while no significant association was observed in women. The Agent further identified key mediating pathways (eGFR reduction and serum creatinine metabolism disorders) and confounding factors (age, gender, BMI) via DAG visualization (Figure 8), verifying the biological plausibility of the causal relationship—thyroid dysfunction may affect renal hemodynamics and filtration function, thereby promoting CKD development^27,28^.

**Figure 8:**
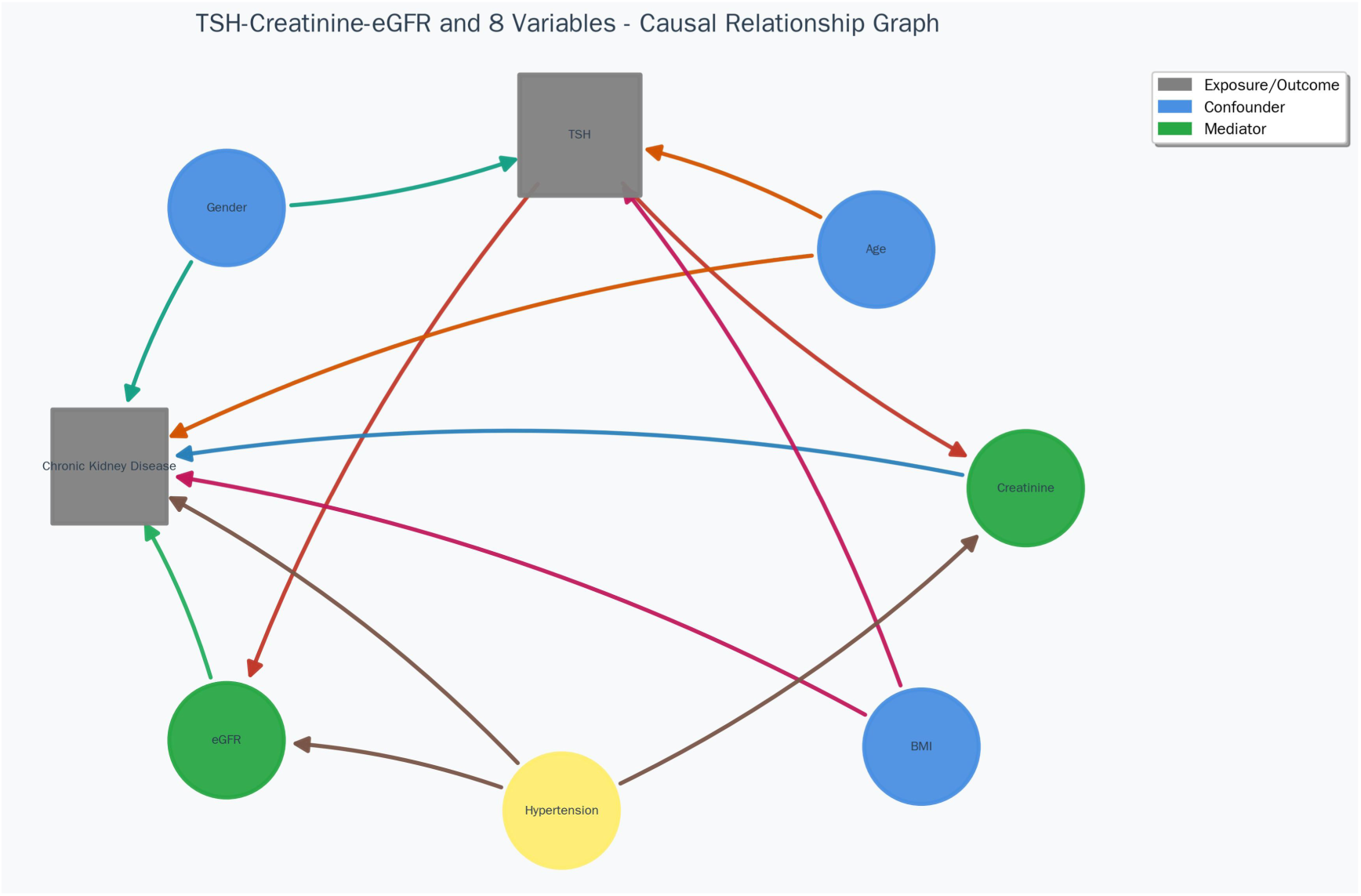
Directed Acyclic Graph (DAG) Illustrating Causal Relationships Between TSH and Chronic Kidney Disease (CKD) This figure presents a DAG visualizing the causal pathways, confounders, and mediating mechanisms between thyroid stimulating hormone (TSH) and chronic kidney disease (CKD). Consistent with the unified visual coding, gray rectangles represent the exposure variable (TSH) and outcome variable (CKD), blue circles denote confounders, and green circles indicate mediator variables. Specifically, the confounders include age, gender, and body mass index (BMI), which are associated with both TSH levels and CKD incidence; the mediators are estimated glomerular filtration rate (eGFR) and serum creatinine, which lie on the causal pathway linking TSH to CKD. Directed edges in the graph show that TSH affects CKD through two mediating pathways (TSH → eGFR reduction → CKD and TSH → serum creatinine metabolism disorder → CKD), while confounders independently influence both the exposure and outcome. This DAG provides a clear visual framework for identifying the minimal sufficient adjustment set (age, gender, BMI) and clarifying the biological mechanism of TSH-related CKD development, supporting unbiased causal effect estimation and clinical decision-making.

From a practical application perspective, the Agent’s results provide clear clinical guidance: first, regular TSH monitoring should be prioritized for middle-aged and elderly men with high BMI, as this population has a higher risk of TSH-related CKD; second, for CKD patients with elevated TSH, targeted thyroid function intervention may reverse part of the renal function impairment, given the reversible nature of thyroid dysfunction in CKD^28^; third, the gender-specific effect reminds clinicians to avoid a one-size-fits-all approach when assessing TSH-related CKD risk, improving the accuracy of risk stratification.

## Discussion

The EpiCausalX Agent, developed in this study as an AI tool specifically tailored for epidemiological causal inference, offers extensive application potential. Its core functional modules—encompassing multi-knowledge base literature retrieval, Directed Acyclic Graph (DAG) visualization, clinical measurement standard query, and causal inference analysis—effectively cover multiple key stages of epidemiological research. Observational studies stand as a primary method in epidemiology, yet causal inference remains a central challenge, and the Agent addresses this with distinct advantages: it automatically identifies potential confounders and generates DAGs, aligning with medical literature that emphasizes the pivotal role of DAGs in clinical research and confounding control^14^; it determines minimal sufficient adjustment sets through DAG analysis, consistent with findings from studies on measurement error and information bias that highlight DAGs’ value in mapping epidemiological concepts ^29^; it provides evidence-based recommendations for selecting causal inference methods, drawing on the theoretical framework of research unifying instrumental variable and inverse probability weighting approaches^30^; and it assesses the impact of unmeasured confounding in sensitivity analysis, supported by established methods for causal inference in observational studies^31^.

Beyond observational studies, the Agent is applicable to diverse clinical research designs, including clinical trials, cohort studies, case-control studies, and cross-sectional studies. In clinical trials, it facilitates baseline variable selection, subgroup analysis, and mediation analysis to enhance efficiency and reduce bias; in cohort studies, it streamlines exposure definition, outcome measurement, and confounding control through automated, standardized workflows; in case-control studies, it aids matching factor selection and bias assessment to improve research quality; and in cross-sectional studies, it supports association analysis and causal hypothesis testing for systematic design. This end-to-end support, spanning from study design to data analysis, aligns with the Agent’s design philosophy of integrating multi-knowledge base retrieval and causal inference, as well as research exploring machine learning’s potential in clinical practice^32^.

Additionally, the Agent holds substantial value in epidemiological education: it enhances teaching effectiveness through visualized DAG generation, interactive learning with real-time DAGs and analytical recommendations for user-input scenarios, case studies grounded in real literature, and standardized training with unified terminology—responding to strategies for integrating AI into medical education and the proven potential of generative AI in teaching^33,34^.

Compared with existing general LLMs (e.g., DeepSeek, GPT-4), the EpiCausalX Agent exhibits notable differences and advantages in epidemiological causal inference, particularly in specialization, reliability, and verifiability. While general large models excel in natural language processing and general question-answering, they face limitations in deep specialized applications. Table 2 summarizes these key distinctions. Thirunavukarasu et al. (2023) noted that general large language models often lack sufficient accuracy and reliability in specific clinical tasks—a limitation the EpiCausalX Agent overcomes by integrating specialized knowledge bases and domain-specific tools^35^. Furthermore, general large models suffer from poor output verifiability due to complex training data and black-box characteristics, whereas the EpiCausalX Agent enhances reliability through four key mechanisms: traceable knowledge sources with complete citation details (including PMID, journal, and authors), standardized methods aligned with epidemiological guidelines, reproducible results from consistent inputs, and multi-source cross-verification of critical conclusions—meeting the emphasis on interpretability and verifiability in medical AI applications.

**Table 2.**
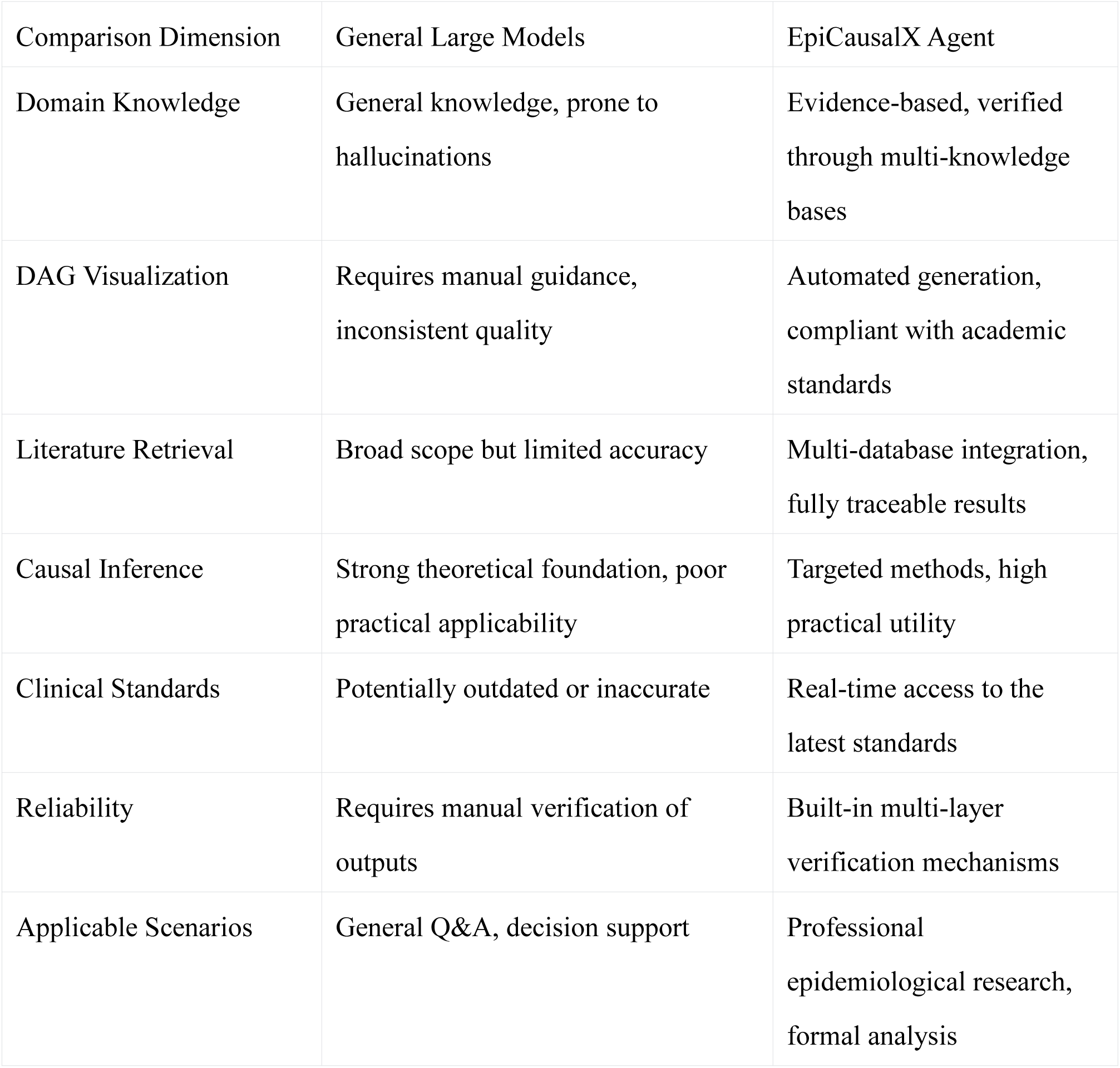
Key Differences Between General Large Language and EpiCausalX Agent Models.

Compared with existing methods, the EpiCausalX Agent boasts significant strengths: it achieves end-to-end automation to minimize manual intervention, supported by research highlighting the value of automation in medical machine learning applications^36^; it is specialized in epidemiological causal inference, aligning with DAGs’ central role in clinical research^14^; its modular design enables seamless function extension, as demonstrated in studies on machine learning-based clinical decision support^37^; its user-friendly interface lowers entry barriers, validated by the study’s usability results; all knowledge sources are traceable via multi-knowledge base integration; and it adheres to epidemiological standard methods, complying with guidelines for comparative effectiveness research^38^. The Agent still has scope for improvement. It currently focuses on epidemiological causal inference, with potential expansion to other medical domains, and primarily supports Chinese and English, with plans to add more languages; in addition, its retrieval algorithm needs optimization to boost real-time performance. These limitations are addressed in its future development roadmap, which also responds to the core challenges of AI applications in epidemiology, such as model interpretability and data privacy protection.

Building on current achievements and limitations, the EpiCausalX Agent’s future development will focus on technical advancement, application expansion, expert validation, and continuous optimization. Technically, it will pursue multimodal integration of diverse data types (text, images, time series), real-time data stream processing for dynamic causal inference, multi-user collaboration features, and cloud deployment to reduce usage barriers—aligning with emerging directions of AI in medical fields^39^. Application-wise, it will expand to public health policy development (supporting intervention effect evaluation), integration with clinical decision support systems (providing tools for clinicians), specialized epidemiological education versions, and data sharing platforms—leveraging AI’s significant potential in public health and medical education^40^. To ensure scientific rigor, the Agent will undergo systematic expert review involving renowned domestic and international experts, functional validation in real-world research scenarios, large-scale user testing, case comparisons with manual analysis, and peer review via academic publication, which aligns with the pivotal role of expert validation in the development of AI medical tools. A continuous optimization mechanism will be established, including monthly knowledge base updates, quarterly model optimization, ongoing feature expansion and performance improvement, sustained user support, and biannual security audits; this mechanism is underpinned by research highlighting the necessity of continuous iteration in healthcare machine learning applications^41^. After initial validation, the Agent will be released as an open-source project to facilitate community-driven optimization, and a detailed roadmap will be formulated to ensure that functional enhancements do not undermine the system’s stability and reliability.

In conclusion, the EpiCausalX Agent automates epidemiological causal inference workflows, with broad applications in observational/clinical research and epidemiological education. Excelling in specialization, reliability, and verifiability, it addresses general large language models’ professional research limitations. As an AI-epidemiology integration, it boosts efficiency via automation and standardized processes, poised to become an indispensable tool for researchers, supporting public health and clinical practice with evidence-based insights.

## Data Availability

All data produced in the present study are available upon reasonable request to the authors

